# Telemedicine in Eye Care During the COVID-19 Pandemic: A Review of Patient & Physician Perspectives

**DOI:** 10.1101/2024.10.25.24316160

**Authors:** Christina Perjuhi Halajyan, Jonathan Thomas, Benjamin Xu, Jeffrey Gluckstein, Xuejuan Jiang

## Abstract

**Purpose:** There has been an increase in the adoption of telemedicine during the COVID-19 pandemic. This review used systematic search and review criteria to assess the literature on patient and physician perspectives toward telemedicine for vision care during the pandemic.

**Methods:** We conducted a comprehensive search on PubMed, Embase, and Scopus using relevant MeSH terms to identify peer-reviewed studies examining telemedicine use in eye care during the pandemic. The search strategy encompassed three key concepts: COVID-19 or pandemic, telehealth or telemedicine, and eye care. Further screening of references and similar articles was conducted to identify additional relevant studies.

**Results:** We identified 24 relevant studies published between 2020 and 2022. Of these, 15 focused on patients’ perspectives, while 12 explored physicians’ perspectives. Predominantly cross-sectional in design, these studies were mainly conducted during the initial wave of the pandemic (March 2020 to June 2020), primarily in urban locations and hospital settings. Patients were satisfied with telemedicine and considered it equally effective to in-person visits. Patients believed telemedicine was convenient, improved eye care access, and a beneficial triage tool. Physicians acknowledged telemedicine’s convenience for follow-up assessment and its ability to expand the capacity for emergency cases. However, both patients and physicians voiced concerns about the absence of ancillary examination and technological challenges.

**Conclusion:** Our review highlights the positive impact of telemedicine in eye care during the pandemic. Nonetheless, most studies were limited in sample size. They did not delve into potential disparities based on race/ethnicity, socioeconomic status, and geographic location, factors that could influence patient attitudes toward telemedicine. Further research is warranted to validate the findings from our selected studies and explore factors that influence the implementation of telemedicine, particularly across various eye care subspecialties.

## INTRODUCTION

Before the onset of the COVID-19 pandemic, telemedicine had been underutilized due to challenges with limited access,^1^ low reimbursement,^1^ interstate licensing,^1^ privacy issues,^1,2^ workflow integration,^3^ and concerns regarding efficacy.^3^ However, it has been recognized that telemedicine can improve access to healthcare, lower costs, reduce travel and wait times, facilitate evaluations of patients with limited mobility or communicable diseases, and provide quality care comparable to in-person visits.^4^ Additionally, telemedicine is considered an effective triage tool to initiate care and determine the need for in-person visits.^5-8^

As a direct response to the COVID-19 pandemic, telemedicine emerged as the primary, and often the only, method of delivering care, especially early on.^3^ Ophthalmology experienced the most significant decline in in-person patient visits across different medical specialties.^9^ Telemedicine use surged among ophthalmologists during the pandemic, with reports from the Philippines^10^ showing an increase from 53% to 90% and nationwide surveys in the U.S.^11^ revealing a 3.4% to 77.4% increase. Despite this surge, ophthalmology still exhibited relatively lower elective use of telemedicine compared to other specialties of medicine.^12^

After the pandemic, many studies have reported data regarding perspectives toward telemedicine utilization. However, these data need to be systematically evaluated and synthesized to improve the implementation of telemedicine in the future. In this review, we used systematic methods to compile a list of existing literature on patient and physician perspectives (e.g., satisfaction, future use) toward telemedicine in eye care during the pandemic. We also evaluated variations in perspectives by factors such as demographics and care settings, which have been shown to influence personal satisfaction toward telemedicine.^13^ Finally, we identified issues with existing literature and critical knowledge gaps. Our goal is to better understand the benefits of and barriers to telemedicine use in eye care and highlight factors that hinder adequate and efficient eye care delivery. Given the lack of consensus on instruments measuring attitudes and the diverse definitions used across studies, we conducted a narrative analysis to synthesize the data comprehensively and provide a nuanced understanding of the landscape.

## METHODS

The methods and findings of this review followed the Preferred Reporting Items for Systematic Reviews and Meta-Analyses guidelines (PRISMA) as shown in Fig 1 and S1 Figure. The study complied with research ethics regulations at the University of Southern California.

**Fig 1.**
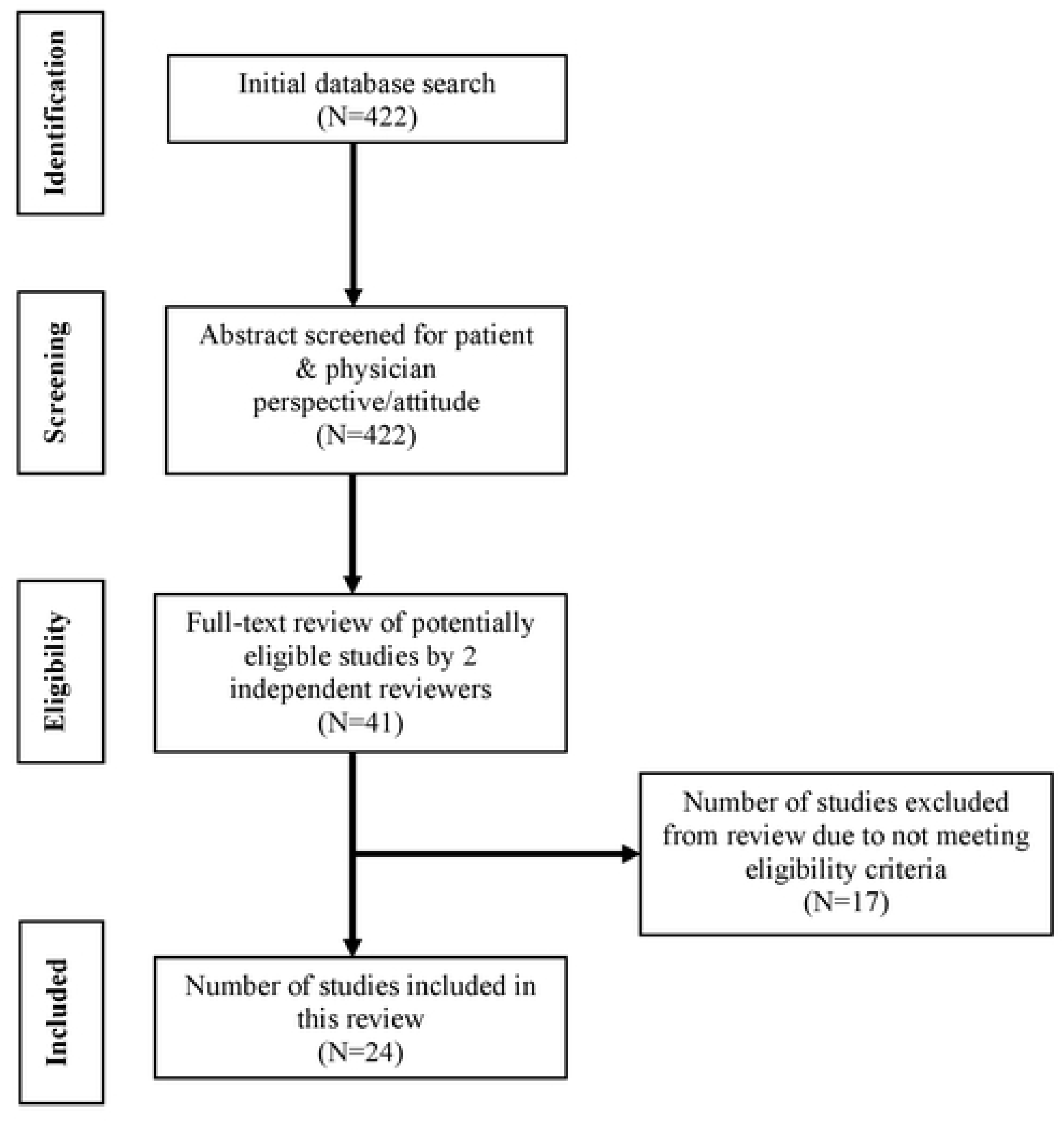
PRISMA Flow Diagram.

### Search Strategy

We searched PubMed, Embase, and Scopus databases until December 2022 to identify peer-reviewed studies that evaluated telemedicine use in eye care during the COVID-19 pandemic, using different combinations of keywords and controlled vocabulary (e.g., MeSH terms). The search terms were a combination of 3 concepts: 1) COVID-19 or pandemic (Keywords: COVID-19, COVID, pandemic) 2) telehealth or telemedicine (keywords: telehealth, telemedicine, teleophthalmology, tele-optometry, virtual health, video visit), and 3) eye care (keywords: ophthalmology, optometry, eye care, vision testing). The detailed search strategy is provided in S2 Appendix. We also searched: 1) the reference lists of all the articles identified, 2) other articles that cited the identified articles, and 3) similar articles indicated by Google Scholar to identify additional studies that may be included in our review. The search strategy was conducted separately by C.P.H. and J.T. Discrepancies between C.P.H. and J.T. were discussed and adjudicated by X.J.

The terms telehealth, telemedicine, and teleophthalmology were used interchangeably across the selected studies. The various communication modalities used in studies included telephone, video visits, e-mail, direct text, and social media messaging. For this review, we used the term telemedicine to encompass the different types of virtual platforms mentioned in the chosen articles. We also retained the language used in those respective studies when referring to specific study findings.

### Eligibility/Exclusion Criteria

We considered only original research articles that had been published in peer-reviewed journals. There were no restrictions on geographic locations or age groups. We included all publications examining direct, synchronous communication modalities between patients and physicians, but excluded ones that only mentioned health monitoring via apps and wearable smart devices. We excluded any article whose primary focus was not telemedicine, including articles related to modeling studies and studies that did not provide immediate and direct benefits for healthcare workers (including medical students and healthcare managers) or patients. Additional studies were excluded if published before March 2020, describing medical specialties other than ophthalmology and/or optometry, unavailable in English, not observing either patient or physician perspectives, and small in sample size (less than 5).

Data were systematically extracted from the identified studies. Information extracted from each study included study design, study site (geographic location and practice type), the study population’s size and characteristics, and telemedicine modality. Our review also focuses on the factors influencing perceptions toward telemedicine and its future use, including age, communication modality preference, differences among subspecialties, practice types, race/ethnicity, and income.

### Outcome Assessment

Patient and physician attitudes toward telemedicine were assessed using satisfaction or other equivalent attitude measures. Patient satisfaction was measured differently across the selected studies. Since the selected studies were heterogeneous in their outcome measurement (e.g., different definitions of satisfaction, 4 vs. 5-point Likert scale for satisfaction), we did not carry out meta-analyses of satisfaction or future use.

Our primary outcomes of interest were patient/physician satisfaction with telemedicine care, defined as responding positively to the original study’s specific definition, and future telemedicine use, defined as patients and physicians that explicitly stated they would use telemedicine post-pandemic. To quantify satisfaction, we cited percentages of patients/physicians who responded positively to the original study’s specific definition. Similarly, the proportion of participants who explicitly stated they would use telemedicine even after the pandemic resolves was used to measure future use. We also estimated 95% confidence intervals for the proportions of satisfaction and future use based on each study’s reported sample size.

### Assessment of Study Quality

We assessed the risk of bias in the included studies with a version of the National Heart, Lung, and Blood Institute (NHLBL) Quality Assessment Tool for Observational Cohort and Cross-Sectional Studies [https://www.nhlbi.nih.gov/health-topics/study-quality-assessment-tools],^14^ modified to better tailor the analysis to our studies. Questions excluded in risk of bias assessment include those relating to the timing or levels of exposure, blinding of outcome assessors, and confounding variables. The modified version utilized in this study can be found in S3 Figure.

## RESULTS

A total of 24 articles (Table 1) were included in our analysis. These studies examined either patients’ or physicians’ attitudes on the implementation of telemedicine in eye care with the emergence of the COVID-19 pandemic: 12 (50%) from patients’ perspective only, 9 (38%) from physicians’ perspective only, and 3 (12%) analyzing both. 12 (50%) of the studies were conducted in the U.S. and 5 (21%) in the U.K. Most studies were conducted during the initial wave of the pandemic from March 2020 to June 2020 (67%, Fig 2), in urban locations (50%), and with a cross-sectional study design (88%). Studies were carried out in comprehensive care centers/hospitals (n=14, 58%), private practices (n=5, 21%) or a combination of settings (n=5, 21%).

**Fig 2.**
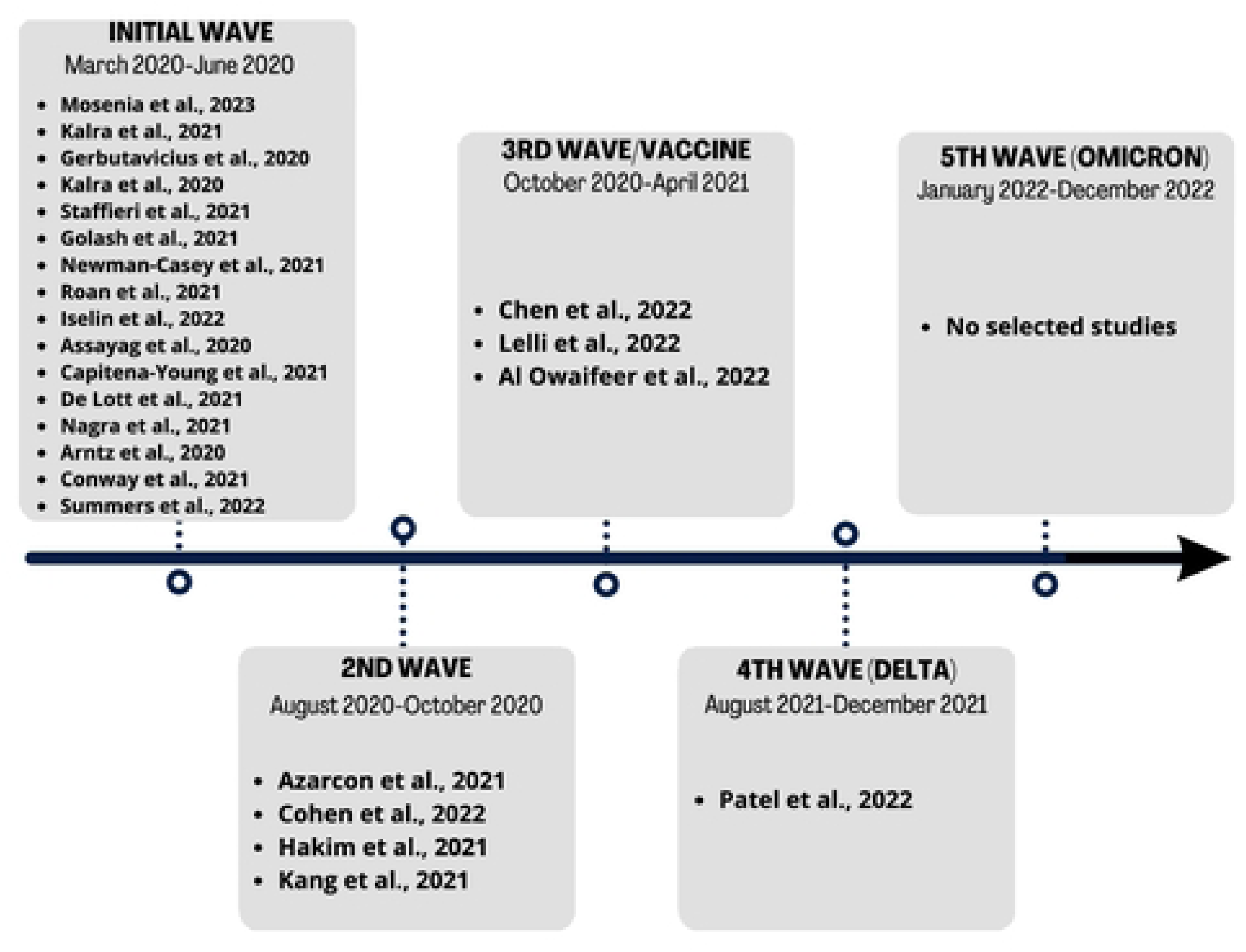
Timeline Illustrating the Chronological Order of the Included Studies During the COVID-19 Pandemic.

**Table 1:** Key Features of Studies Included in This Review.

| Study | Participants | Geographic Location<br>(urban or rural) | Practice Type | Study Design | Modality Used | Preferred Modality |
| --- | --- | --- | --- | --- | --- | --- |
| Mosenia et al., 2023 | 123 patients<br>(unknown) | San Francisco, USA (urban) | Tertiary Academic<br>Medical Center | Retrospective, cross-sectional | Video, phone,<br>hybrid<br>(asynchronous +<br>synchronous) | N/A |
| Kalra et al., 2021 | 56 patients (adult) | Pennsylvania, USA (urban) | Tertiary Academic<br>Medical Center | Retrospective, pre-<br>and post-COVID<br>analysis | Telephone and<br>virtual (video &<br>eConsult) | Virtual |
| Gerbutavicius et al.,<br>2020 | 29 patients (adult) | Germany (N/A) | Ophthalmology<br>Practice | Cross-sectional | Video Consultation | N/A |
| Kalra et al., 2020 | 92 patients (adult) | Pennsylvania, USA (urban) | University Hospital | Retrospective, cross-sectional | Video | N/A |
| Staffieri et al., 2021 | 89 patients<br>(pediatric) | Australia (N/A) | Private Pediatric<br>Ophthalmology<br>Practice | Cross-sectional | “Telehealth” | N/A |
| Golash et al., 2021 | 120 patients (adult)<br>(80 phone, 40<br>video) | Maidstone District of Kent,<br>England (rural) | General Hospital | Cross-sectional | Telephone & video | Video |
| Newman-Casey et al.,<br>2021 | 1,720 patients<br>(adult) | Michigan, USA (urban) | Tertiary,<br>Multispecialty Care<br>Practice | Cross-sectional | Telephone & video | Video |
| Roan et al., 2021 | 211 patients<br>(virtual)<br>307 patients (in-person) | Ohio, USA (urban) | Comprehensive<br>Care Center | Retrospective,<br>observational | Video-<br>conferencing,<br>phone-call, in-person | N/A |
| Iselin et al., 2022 | 69 patients (adult) | Dublin, Ireland (urban) | Tertiary, single<br>center | Prospective, cross-sectional | Telephone, video,<br>email | N/A |
| Hakim et al., 2021 | 51 patients (adult) | Cheshire, UK (rural) | General Hospital | Cross-sectional | Telephone & video | N/A |
| Chen et al., 2022 | 252 patients (adult) | New York, USA (urban) | Tertiary Care<br>Center | Retrospective, single-<br>center, cross-sectional | Synchronous video<br>visits | N/A |
| Patel et al., 2022 | 103 patients (adult) | Illinois, USA (rural) | Multi-provider,<br>Retina-only<br>Practice | Cross-sectional | "Telemedicine" | 68.7%<br>preferred<br>in-person |
| Assayag et al., 2020 | 70 physicians | Multiple countries; majority<br>USA, Israel, India (N/A) | Hospitals, Private<br>and Community<br>Clinics | Cross-sectional | "Telemedicine" | N/A |
| Capitena Young et al.,<br>2021 | 117 physicians | Nationwide, USA (N/A) | Academic Center,<br>Private Practice,<br>Group Practice,<br>Hospital - multi-<br>type | Cross-sectional | Telephone, video,<br>email | Telephone |
| De Lott et al., 2021 | 88 physicians (73<br>ophthalmologists,<br>15 optometrists) | Michigan, USA (urban) | Tertiary Referral<br>Center | Cross-sectional | Telephone,<br>interprofessional<br>consultations,<br>video | N/A |
| Nagra et al., 2021 | 1,250 physicians<br>(optometrists) | England, Scotland, Wales,<br>Northern Ireland (N/A) | Primary Care<br>Optometry<br>Practices | Cross-sectional | Telephone only,<br>telephone/video | N/A |
| Azarcon et al., 2021 | 327 physicians | Philippines (N/A) | Private Practice | Prospective,<br>descriptive | Email, telephone,<br>short message<br>service, social<br>media messaging,<br>video | Social<br>Media<br>Messaging |
| Cohen et al., 2022 | 361 physicians | Nationwide, USA (N/A) | Retina Only Private<br>Practice,<br>Multispecialty,<br>Solo, University<br>based | Cross-sectional | Direct audio phone<br>call, video, direct<br>text messaging | N/A |
| Kang et al., 2021 | 45 physicians | England, Ireland, Scotland<br>(N/A) | Teaching Hospitals,<br>District General<br>Hospitals, Tertiary<br>Ophthalmic Centre,<br>Private Practice | Cross-sectional | Telephone, video,<br>asynchronous<br>methods | Video |
| Lelli et al., 2022 | 192 physicians | Multiple countries, majority<br>USA (N/A) | Self-employed,<br>University, Private | Cross-sectional | Real-time video<br>visit, audio | N/A |

|  |  |  | Practice, Hospital,<br>Other |  | only/telephone<br>visit, store-and-<br>forward<br>imaging/video,<br>technician-based<br>in-person |  |
| --- | --- | --- | --- | --- | --- | --- |
| Al Owaifeer et al.,<br>2022 | 71 physicians | Saudi Arabia (urban) | Tertiary Eye Care<br>Center | Descriptive, cross-sectional | Telephone, video,<br>email | N/A |
| Arntz et al., 2020 | 91 patients (adults)<br>12 physicians | Chile (urban) | Private Healthcare<br>Network | Cross-sectional | Video | N/A |
| Conway et al., 2021 | 159 patients<br>(adults)<br>157 physicians | New York, USA (urban) | Academic Tertiary<br>Care Centers | Multi-center, cross-sectional | Video | N/A |
| Summers et al., 2022 | 143 patients<br>(general)<br>34 physicians | Oregon, USA (urban) | Academic Medical<br>Center/Tertiary<br>Referral Center | Retrospective, cross-sectional | In-office,<br>synchronous<br>audiovisual, audio<br>only, imaging only,<br>support staff only | N/A |

The selected studies included patients and physicians from different subspecialties of eye care, including comprehensive, oculoplastic, neuro-ophthalmology, cornea, glaucoma, pediatrics, and retina. There were disproportionately more studies (17%) focusing solely on telemedicine among the oculoplastic subspecialty than other subspecialties.^15-18^ Utilization of telemedicine in eye care seemed to differ by subspecialties; however, findings were not consistent across the studies. For example, a retrospective study^19^ reported telemedicine being utilized the most by patients in general ophthalmology, cornea/cataract, and retina. Another study^20^ conducting synchronous video visits during the pandemic found the volume of telemedicine visits for patients was highest among oculoplastic surgery (42.9% of all visits), followed by neuro-ophthalmology (17.0%) and cornea (14.2%).

Most of the chosen studies mainly describe the patient-physician encounter without presenting possible interventions. A few studies^5,10,19,21,22^ did report that a common reason for a telemedicine visit was for medication prescriptions. Two studies^17,23^ also mentioned that some of their virtual visits consisted of follow up evaluations to treatments such as intravitreal injections but did not specify where or how these treatments were done.

Our review was organized into four parts: 1) patients’ perspective; 2) physicians’ perspective; 3) common factors that influenced both patients’ and physicians’ perspectives; and 4) evaluation of the quality of the selected studies.

## Patient Perspective

### Patient Satisfaction

Common indicators of patient satisfaction and preference for telemedicine include recurrent virtual visits,^20^ seamless use of technology,^6,20,23,24^ pleased with the service provided during the visit,^6-8,23,24^ instructions provided prior to the visit were clear and easy to understand^6,15,25^ and not feeling rushed and that concerns were listened to.^12,15^ Lastly, if participants found the visit convenient, timesaving,^5-8,15,20,24^ helpful,^19,21^ and safe^7,15,19,21^ also implied being content with the care delivery.

Fig 3 presents the patient perspective studies with a direct measure of the proportion of patients who were satisfied. Four studies^5,22,24,26^ reported other quantifications of attitude and, therefore, were not included in Fig 3. Most studies in Fig 3 reported an overall patient satisfaction of 65% or greater. Among the 4 studies not included in Fig 3, Patel et al.^26^ found a generally neutral attitude toward telemedicine among patients with retinal disease, with 18.4% reporting positive attitudes. The definition of positive attitudes as having ≥4 on a 5-point Likert scale for all 14 questions in six domains likely accounts for the low level of positivity.^26^ On the other hand, Summers et al.^24^ found that patients who participated in telemedicine visits were more likely to recommend the institution or physician that conducted the visit than those who had in-office visits (recommended institution: 81.4 vs. 79.6; recommended provider: 92.3 vs. 86.4, respectively). Unfortunately, no statistical testing of these differences was reported. Kalra et al.^5^ found that patients rated their overall experience with video visits as favorable compared to their regular consult visits. Roan et al.^22^ found comparable patient satisfaction scores for virtual and in-person visits.

**Fig 3.**
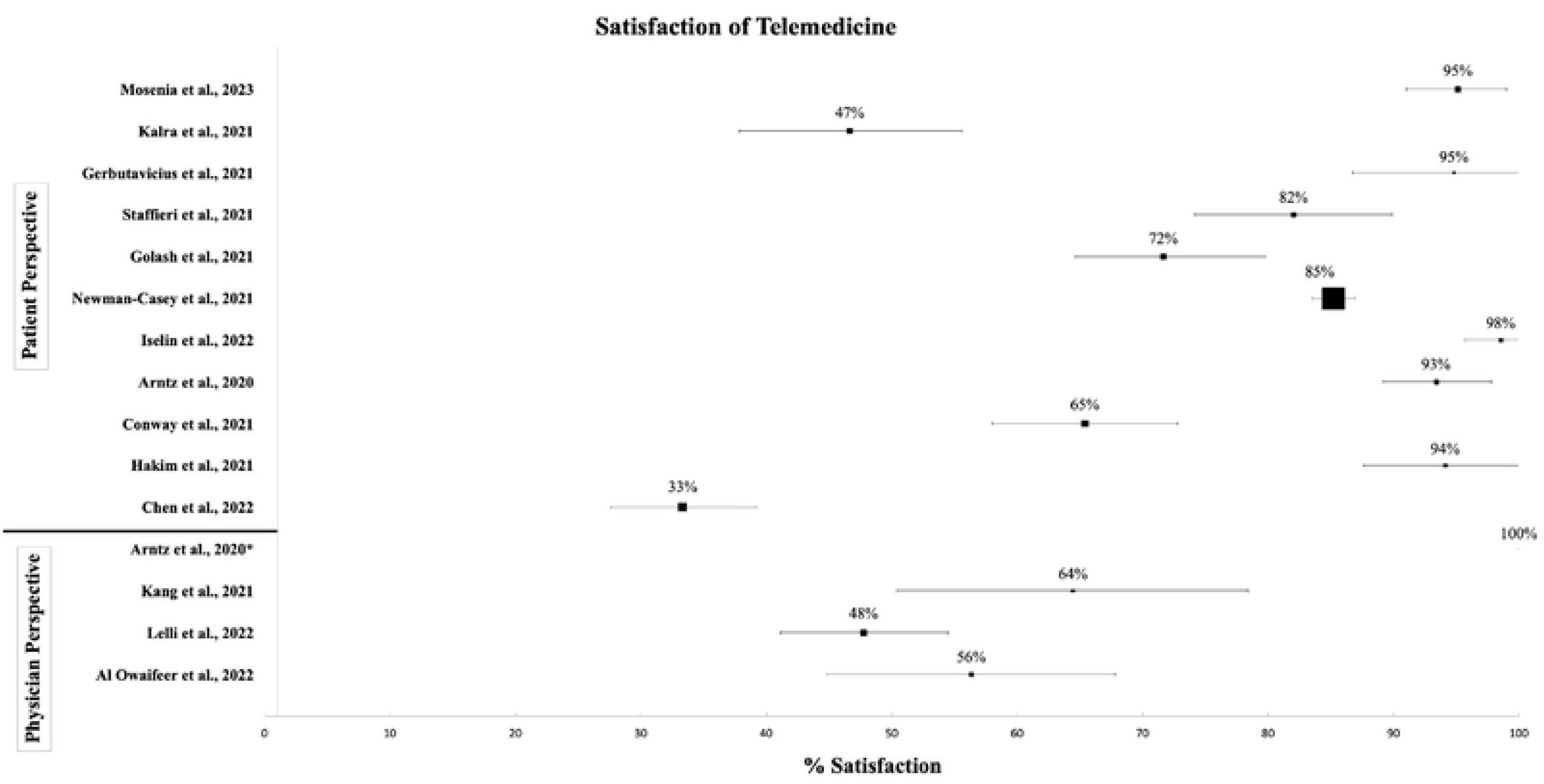
Forest Plot of Patient- and Physician-Satisfaction of Telemedicine in Included Studies. Note: bars indicate the 95% confidence interval, and the size of the box corresponds to the relative size of the study sample *= This study’s sample size is n=12 with a satisfaction event of n=12/12

### Patient Benefits

Respondents noted that telemedicine was convenient^5-8,15,20^ and timesaving in terms of travel, parking, gas expenses, and arranging time off work and childcare (Fig 4).^5,6,15,24,27^ In one study,^27^ participants noted that the average waiting time decreased from 43 minutes for an in-person visit to 4 minutes with the virtual clinic. Telemedicine was also convenient for patients dependent on others, which accounts for 34% of the patient population in one study.^15^ Nevertheless, Kalra et al.^19^ revealed that the role of “teleophthalmology” was restricted to triage, early management, counseling, and follow-up with stable patients.

**Fig 4.**
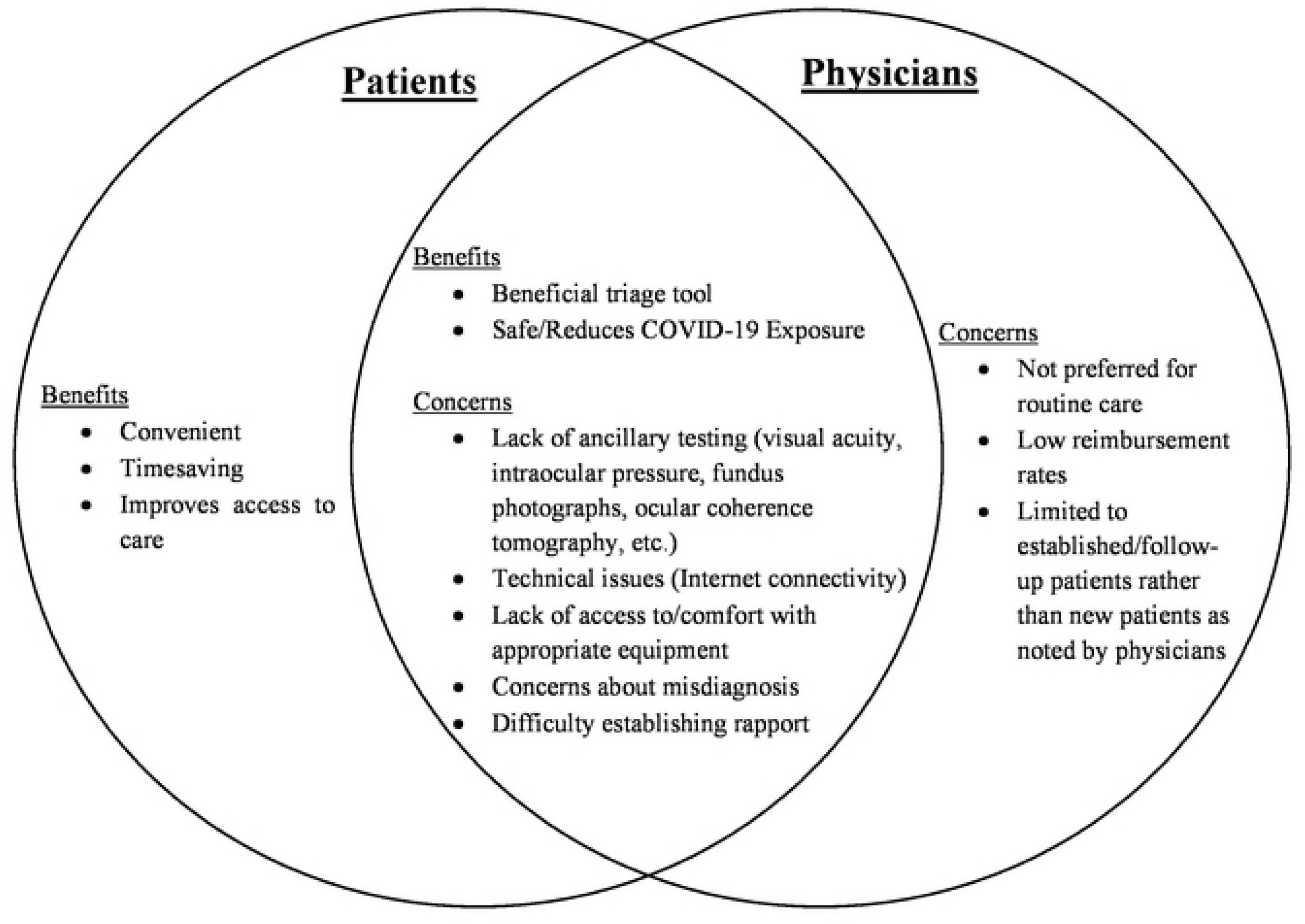
Summary of Patient’s and Physician’s Views of the Benefits and Concerns of Telemedicine for Eye Care.

As a beneficial triage tool, Chen et al.^20^ found that 65% of patients had their issues addressed over the virtual visits. Newman-Casey et al.^7^ found that only 6.8% of virtual visits resulted in an in-person visit within 2 weeks. These findings support telemedicine as an effective triaging system for eye care.

### Patient Concerns

Common concerns prevalent across studies (Fig 4) include worries over lack of ancillary examination,^6-8,12, 25^ technological challenges (e.g., internet connectivity, lighting, suitable space, phone positioning, and unsuccessful video links),^6,15,23-25^ and unclear login instructions.^6,15^ Some patients preferred using a platform they had already gotten accustomed to (such as Zoom) rather than using one integrated into their electronic medical record.^20,25^ Patients in another study ^7^ also expressed concerns about difficulties establishing rapport with their physician during a virtual visit.

### Willingness to Use Telemedicine in the Future

More than half of the participants favored telemedicine use in the future (Fig 5).^5,6,21,26,27^ Patients reported that in the absence of a video visit option, they would have delayed seeking care during the pandemic.^5,8^ Likewise, Staffieri et al.^6^ reported that 71.9% of parents of pediatric patients would consider telemedicine visits for their kids in the future. Patel et al.,^26^ which reported that most patients with retinal disease had a neutral attitude toward telemedicine, also found that among patients with prior telemedicine experience, 66.7% of patients would use telemedicine in the future. These findings suggest that patients were satisfied enough with eye care delivery through telemedicine and would consider it over in-person visits despite their challenges with telemedicine during the pandemic.

**Fig 5.**
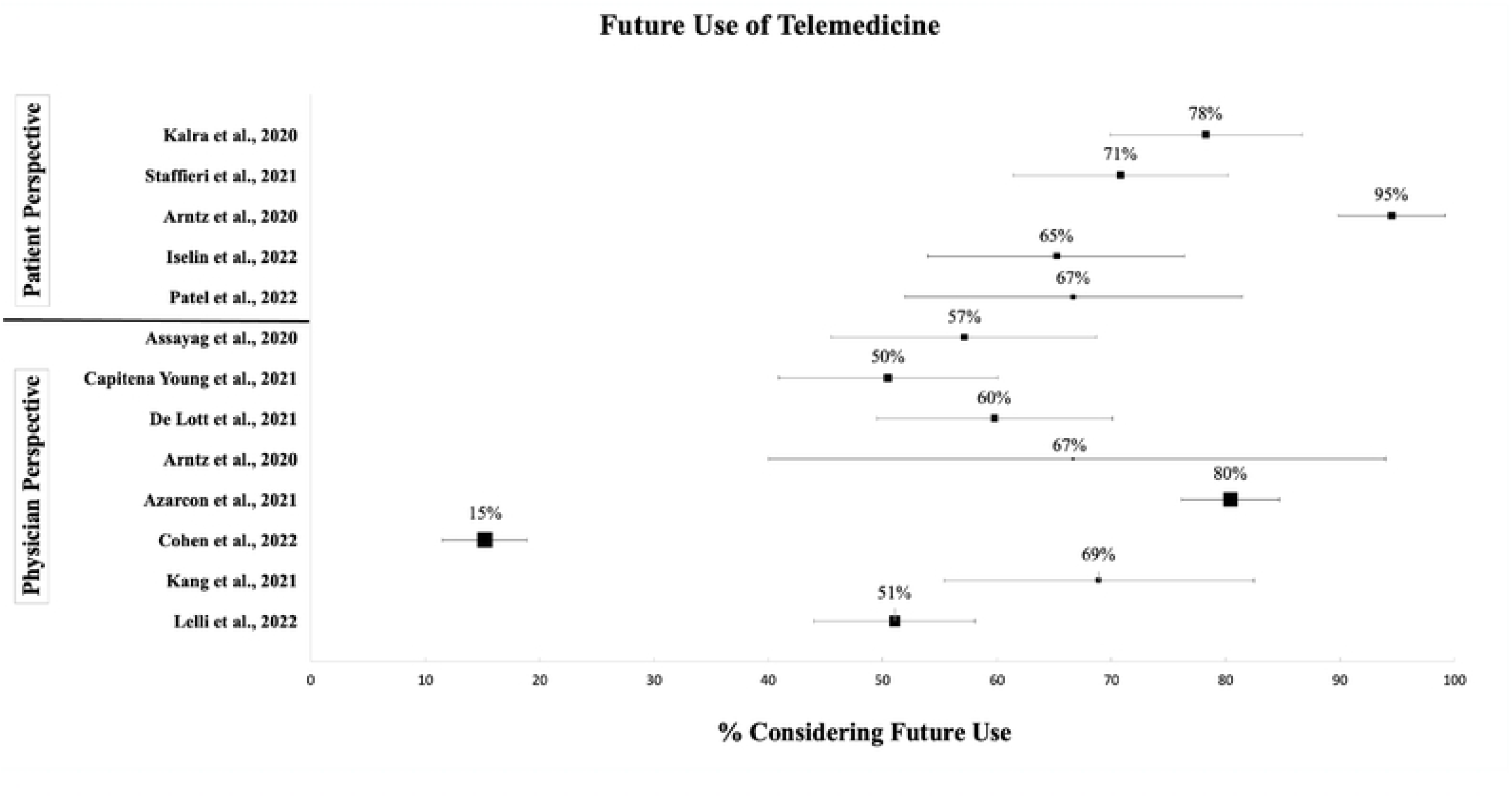
Forest Plot of Patient’s and Physician’s View On Future Use of Telemedicine in Included Studies. Note: bars indicate the 95% confidence interval, and the size of the box corresponds to the relative size of the study sample

## Physician Perspectives

### Physician Satisfaction

Parallel with the patient studies, the physician perspective studies used distinct measurements to gauge attitudes toward telemedicine, including satisfaction,^16,17,21,28^ confidence^10,29^ or comfort^30^ level in utilizing telemedicine, ease of implementation,^11^ ability to gather information for an accurate diagnosis,^25^ and effectiveness.^18,24^ The chosen physician perspective studies include responses from eye care providers in different subspecialties, ranging from general ophthalmologists/optometrists to retina, cornea, and glaucoma specialists.

Fig 3 presents the four studies that measured physicians’ satisfaction levels directly. The reported satisfaction ranged from 48% to 100%. The eight-physician perspective studies^10,11,18,24,25,29-31^ not included in Fig 3 evaluated attitudes toward telemedicine,^24^ its overall usefulness^10,11,18,25,31^ as well as the confidence^29^ and comfort^30^ levels in using telemedicine, instead of overall satisfaction. De Lott et al.^29^ measured confidence levels and found that 66.2% of ophthalmologists and 71.4% of optometrists felt “somewhat confident” in using telemedicine. Nagra et al.^30^ evaluated only optometrists’ comfort level and reported that 83% were “very/moderately” comfortable performing remote consultations. Three studies^10,11,25^ explored whether telemedicine allowed physicians to gather enough information for an accurate diagnosis. Conway et al.^25^ surveyed neuro-ophthalmologists, and 87% of them stated that the examinations they performed provided enough information for medical decision-making. Azarcon et al.^10^ surveyed ophthalmologists in the Philippines and found they were confident with diagnosing gross eye conditions but less confident in diagnosing posterior pole conditions and orbital fractures through telemedicine. After a nationwide survey of eye care providers, Capitena Young et al.^11^ found that 40% of physicians thought the implementation of telemedicine was “somewhat difficult”/ “very difficult,” and 87.5% felt the main reason for the difficulty was related to improper examinations and testing.^11^ Although none of the responding physicians felt that telemedicine was “very reliable,” 50% agreed that virtual evaluations such as those of visual acuity were “somewhat reliable.”^11^

We noted that clinicians had different perspectives on telemedicine depending on the subspecialty. Assayag et al.^18^ surveyed 70 oculoplastic surgeons from 8 countries, and 67.1% of surgeons considered telemedicine an effective tool. On the other hand, Cohen et al.,^31^ which surveyed vitreoretinal specialists, reported that 62.3% of specialists did not think the current form of telemedicine was acceptable for conducting examinations in their specialty. Summers et al.^24^ surveyed providers in an academic pediatric ophthalmology practice and found that providers had more positive attitudes toward telemedicine than staff. Regardless, most responding physicians disagreed with the statement, “I prefer having telehealth visits over clinic visits,” indicating that this group of physicians does not view telemedicine as a valid substitute for traditional eye care delivery.^24^

### Physician Benefits

In general, physicians, both ophthalmologists and optometrists, supported telemedicine use. Similar to patients, physicians also acknowledged (Fig 4) that telemedicine is a convenient tool for assessing follow-up patients,^16,17,28^ for triaging,^16,21^ for continuity of care,^24^ and is helpful in expanding the capacity to see emergency cases in person.^30^ Moreover, it was noted that physicians with more experience practicing telemedicine had higher overall satisfaction scores^28^ and felt more confident^29^ carrying out eye care services through telemedicine, which in turn resulted in higher utilization rates.

### Physician Concerns

Physicians had some concerns regarding telemedicine (Fig 4), some overlapping with those expressed by patients. The primary concern was the lack of testing and imaging abilities during a remote consultation,^10,11,16,25,28,30,31^ which could lead to misdiagnoses.^28,31^ As a consequence, some eye care providers found telemedicine unreliable.^11,28,30,31^ This was especially true for vitreoretinal specialists, among whom 62.3% indicated that the current form of telemedicine was not acceptable for the issues commonly seen in their specialty.^31^ Still, most physicians believed telemedicine visits could be a safe replacement for standard in-person visits if remote fundus imaging were available.^31^ Comparably, Conway et al.^25^ suggested that telemedicine is limited to and mainly beneficial for external examinations and conditions that can be evaluated based on history and for patients who have had previous ancillary testing.

Physicians also reported encountering technological issues, such as lacking access to technology for telemedicine visits or not feeling comfortable using the technology.^10,16,17,25,28^ In addition, many physicians thought that the care they delivered through telemedicine depended on a patient’s level of preparation^25^ and experience with technology.^28^ Other challenges reported by responding physicians include anxieties regarding telemedicine training,^30^ ambiguous guidelines provided by external regulatory bodies resulting in different interpretations by eye care providers,^30^ low reimbursement rates,^17,31^ and potential liability issues.^31^ One study^17^ even found that 56% of their responding physicians thought telemedicine is not considered timesaving, the opposite of many other selected studies. Physicians also noted difficulties in implementing telemedicine visits. Capitena Young et al.^11^ reported opposition to telemedicine from patients, especially older patients, staff members having difficulty incorporating telemedicine into their practice, difficulty determining which patient was better suited for a telemedicine visit, and trouble gaining consent from patients before the virtual visit.

### Willingness to Use Telemedicine in the Future

8 studies surveyed physicians’ views on future use.^10,11,16-18,21,29,31^ Overall, more than 50% of physicians expressed a desire to use telemedicine in the future (Fig 5). De Lott et al.^29^ reported that 62.1% of physicians felt that telemedicine use was underutilized in eye care, and 59.8% would continue using telemedicine. Similar to the patients’ perspective, a study found that physicians were more likely to implement telemedicine in the future if they had previously used it.^31^ Conversely, Cohen et al.^31^ found that only 15.2% of vitreoretinal specialists were prepared to use telemedicine in the future, due to concerns over inaccurate diagnoses and the technology required to conduct a telemedicine visit. Azarcon et al.^10^ found that although many eye care providers in the Philippines would continue to use telemedicine in the future, the number would be less than the providers using it during the pandemic. Similarly, Capitena Young et al.^11^ reported that although half of the clinicians in their study would use virtual health “routinely” or “sometimes,” a few respondents would only utilize telemedicine for a small portion of their visits. No provider felt most of their visits would be through telemedicine after the pandemic.^11^ In addition, even though most oculoplastic surgeons felt that telemedicine would be a beneficial tool in their practice even after the pandemic,^16-18^ none preferred to use telemedicine alone as their primary form of eye care delivery.^16^

## Factors Affecting Patients’ and/or Physicians’ Use and View of Telemedicine in Eye Care

We identified 5 main themes influencing patients’ or physicians’ views regarding the shift toward telemedicine during the pandemic: age, communication modality preference, eye care subspecialties, practice type, and sociodemographics.

### Differences by patients’ and physicians’ age

A gap exists between younger and older patients/physicians regarding the utilization and views toward telemedicine in eye care;however, the reasons for these differences have not been explored in the selected studies.A random sample of patients from the University of Michigan Kellogg Eye Center^7^ revealed that during the pandemic, patients seen in person were significantly older (mean ± standard deviation of age: 66.8 ±17.3 years) than patients using telephone visits (62.6 ±17.8) or video call visits (59.8±15.0 years). Patel et al.^26^ noted that among patients with retinal diseases, younger age (i.e., <75 years old) was the most important factor associated with a greater preference for telemedicine. While 41.4% of patients younger than 75 years preferred telemedicine over in-person visits, the corresponding figure was only 12.1% among older patients.^26^ Similarly, while 62.5% of oculoplastic patients over the age of 65 requested in-person visits, the corresponding figure was only 18.8% among 25–64-year-old oculoplastic patients.^15^

A similar trend was seen among physicians, with older physicians less likely to use telemedicine than younger physicians. This pattern was consistent across all physician perspective studies. Specifically, younger physicians were more likely to be satisfied^16,28^ and comfortable^30^ with telemedicine use, view telemedicine as an effective tool,^18^ and be more willing to use it post-pandemic.^10,16,18,31^

### Communication Modality Preference for Telemedicine

Many care centers in the selected studies offered different telemedicine modalities, with telephone and video visits being the most common. In general, patients and physicians preferred and scored higher on satisfaction for video consultations over telephone consultations when offered both or all modes of care delivery.^7,11,15-17,19^ On the other hand, a survey of ophthalmologists in the Philippines^10^ found that although the use of most types of communication increased during the pandemic, more than 60% of ophthalmology practices utilized telephone and/or short messaging service (SMS) and social media messaging in addition to virtual consultations. Moreover, a multi-country study of oculoplastic surgeons found that physicians in the U.S. were less likely to use asynchronous forms of telemedicine and more likely to engage in telephone visits than physicians in other countries.^17^ However, in a study of monitoring patients with keratoconus through telemedicine,^27^ patients were satisfied with their virtual care appointment regardless of the modality.

Although not commonly used during the pandemic, a hybrid model was implemented in 4^11,12,22,24^ out of the 24 selected studies (17%). A hybrid care model includes in-person and virtual visits for individuals needing intraocular pressure measurements, retinal photography, etc.^12,22^ In one study,^12^ technicians performed necessary testing at the main ophthalmology facilities, and 6 weeks later, a virtual visit was made so that the patient could discuss the results with a physician through either a phone or video visit.^12^ Similarly, at (OHSU) Casey Eye Institute, adult strabismus patients underwent a hybrid visit with an orthoptist performing sensorimotor examinations in person before a virtual visit was scheduled.^24^

### Comparison Among Subspecialties of Eye Care

Some studies^6,7,20,29^ did not observe a significant difference between satisfaction scores across subspecialties. Physicians in some subspecialties were more satisfied with telemedicine than others. Al Owaifeer et al.^28^ found that satisfaction scores for the retina subspecialty were significantly higher than those for other subspecialties mentioned in their study, including general and pediatric ophthalmology, cornea, and glaucoma.

### Differences Among Practice Types

Some studies^10,17,18^ found that physicians’ views toward telemedicine differed across different practice types. A study in the Philippines^10^ found that ophthalmologists who work in the government setting or private practices were more likely to use teleophthalmology compared to physicians who were self-employed (93.64%, 96.67%, vs. 85.03%, respectively, P=0.027). Another study^17^ found that oculoplastic surgeons working in university-affiliated care centers were more likely to use telemedicine (90%) than self-employed (70%, P<0.01) surgeons. A survey of 70 oculoplastic surgeons also found that 74.3% of those employed by hospitals considered telemedicine an effective tool, while only 56.3% of those working in private practices felt the same way.^29^ However, this difference between practice types was not statistically significant (p=0.12), possibly due to the small sample size (N total=70).^29^ Conversely, Cohen et al.,^31^, surveyed vitreoretinal specialists and did not find that the practice type was a significant indicator of telemedicine use during the pandemic.

### Differences by Race/Ethnicity and Income Level of Patients

Most studies did not explore how sociodemographic factors (such as race/ethnicity and socioeconomic status) impact satisfaction with telemedicine use among patients. Two studies^7,12^ Compared telemedicine use between different racial/ethnic groups. With 97.7% of respondents being non-Hispanic compared to 2.3% being Hispanic, the study found significantly more White patients had in-person visits than minority patients, at 87.3% and 12.6%, respectively (P=0.002).^7^ Mosenia et al. reported that telemedicine with asynchronous testing was mostly utilized by White (46.8%) and Asian (29.4%) patients; but data on whether patients were satisfied with their telemedicine visit was not assessed.^12^ Regarding income level, the study of ophthalmologists practicing in the Philippines found no statistically significant difference in virtual care usage between low-income patients compared to middle and high-income patients.^10^

### Study Quality Assessment

We evaluated 7 aspects of study quality across the selected studies (Fig 6 & S3 Figure). Two common quality issues were low participation rate (Fig 6a) and lack of sample size justification (Fig 6b). 17 studies had a participation rate of less than 50%, and 21 studies did not provide a sample size justification. A low response rate in survey participation could introduce response bias because individuals who were more satisfied with telemedicine may be more likely to participate. Therefore, results from surveys with low responses may be biased towards higher satisfaction or more positive responses. Based on the number of negative qualities on the risk assessment, studies were then classified into a rating of good (1 or no quality concern), fair (2 quality concerns), or poor (3 or more quality concerns) quality. Overall, 11, 9, and 4 studies were deemed good, fair, and poor, respectively. Compared to the other studies, the 4 studies with poor quality did not clearly define satisfaction or future use or did not select and recruit all participants from the same or similar populations.

**Fig 6.**
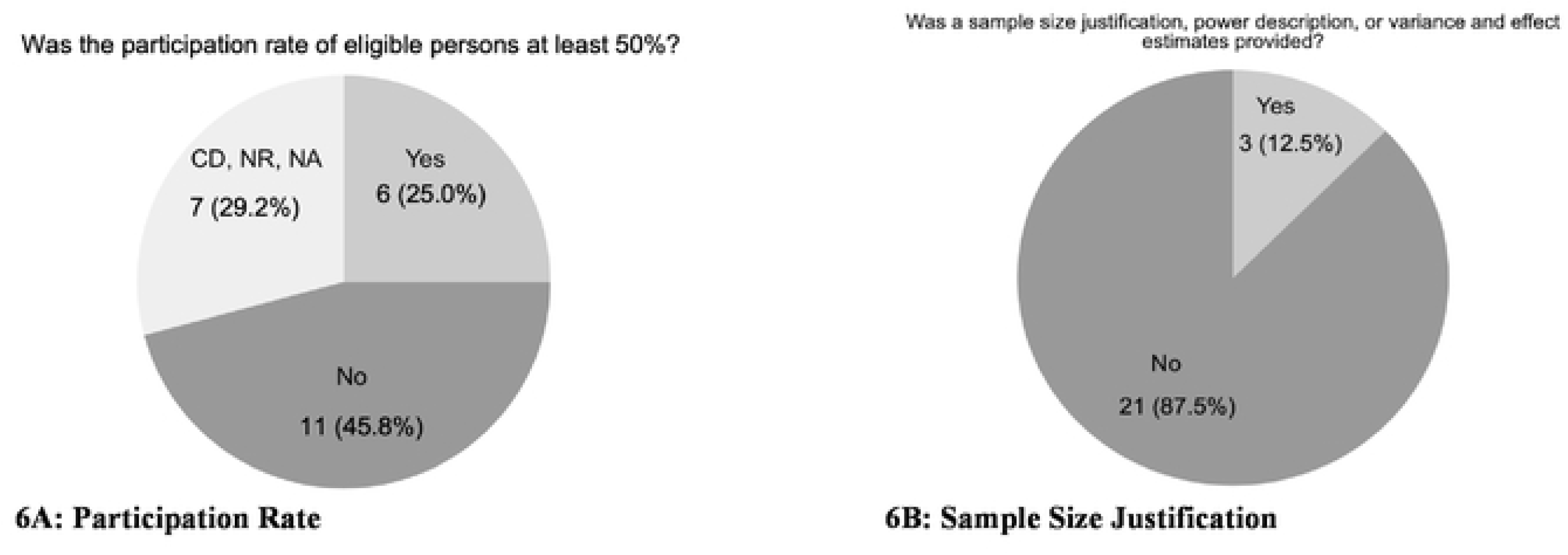
Evaluation of Study Quality – Top Two Study Quality Issues Identified in the Included Studies.

## DISCUSSION

Our review identified 24 studies that provided evidence for telemedicine being acceptable for patients and physicians in eye care during the COVID-19 pandemic. Strengths of these studies include a comprehensive list of eye care subspecialties, comparisons between different communication modalities, representation of various practice types, and a geographically diverse sample. However, it’s important to acknowledge that the identified studies have significant limitations, including low participation rates, inadequate representation of marginalized populations, failure to evaluate the impact of sociodemographic factors, and the use of disparate instruments to measure satisfaction. Despite these limitations, the insights garnered from our review provide a foundation for further refining the implementation of telemedicine within eye care.

Overall, our findings indicate that telemedicine was well received by both patients and physicians in eye care. This aligns with existing literature on other medical specialties such as orthopedics, dermatology, and mental health, which also found that their patients and physicians were satisfied with the remote care provided during the pandemic and preferred video over audio-only visits,^32-38^ similar to patients and physicians in eye care. In particular, both patients and physicians feel telemedicine is helpful as a supplement to traditional in-person visits rather than a complete replacement, acknowledging telemedicine’s strengths in enhancing accessibility and convenience. Although the selected studies demonstrate that telemedicine is emerging as a beneficial platform, there remains ongoing debate regarding its future role.

Eye care providers predominantly used telemedicine for urgent examinations, triaging, adnexal disease, and postoperative care. This is consistent with the fact that eye care relies heavily on specialized imaging and testing that needs to be done in person. Consistently, our findings reveal that telemedicine use did vary by eye care subspecialty with varying levels of reliance on in-person exams. For example, oculoplastic specialists were more inclined to engage in telemedicine visits, particularly video consultations, since follow-up examinations in this subspeciality typically focus on external assessments. Consequently, our selected studies had disproportionately more data from oculoplastic encounters. On the other hand, vitreoretinal specialists, who have major concerns about the absence of virtual testing, reported a lower likelihood of telemedicine use in the future. There were mixed findings regarding telemedicine use in cornea and retinal services compared to other subspecialties. While some studies^5,7,19,20^ found that telemedicine was used more often in these two specialties, others had opposite findings or entirely omitted the cornea and retina specialties in their data. These findings underscore the importance of tailoring telemedicine to the unique demand of different eye care specialties.

One way to overcome the lack of testing is by incorporating asynchronous testing. In contrast to synchronous telemedicine, real-time interaction between the patient and physician through video or telephone, asynchronous telemedicine involves transferring clinical information that the physician will view and report back to the patient with a diagnosis.^1^ In eye care, the asynchronous approach has been utilized to treat and monitor diseases such as diabetic retinopathy. Asynchronous testing was used to augment teleophthalmology visits at ophthalmology clinics of the University of California, San Francisco, mostly for glaucoma, optometry, and cornea encounters.^12^ They found that incorporating asynchronous testing data changed clinical management among 25% of their patients.

More than half of the studies included in this review were conducted during the initial wave of the pandemic from March 2020 to June 2020. Notably, studies conducted in the third wave (Lelli et al., Chen et al., Al Owaifeer et al.)^17,20,28^ showed relatively lower satisfaction than earlier studies. This trend is consistent with the observation from Hakim et al.^8^ that patient satisfaction scores decreased with subsequent virtual visits. The initial wave prompted the implementation of stringent stay-at-home orders, social distancing, and masking guidelines, all aimed at reducing the spread of COVID-19. During this time, telemedicine emerged as a life-saving tool that minimized patient-physician interactions while still providing necessary care.^39^ However, throughout the latter half of 2020 and into 2021, as the fear of the coronavirus waned and vaccines became available in the third wave of the pandemic, patients and physicians were more inclined to “return to normal.” This finding corresponds with studies reporting decreased telemedicine use in their practice during the third wave.^40,41^ Moss et al.^41^ observed that the proportion of neuro-ophthalmologists using video visits reduced from 67% during the initial wave of the pandemic to 50.8% one year into the pandemic. These findings suggest a dynamic evolution in patient and physician perspectives regarding telemedicine and its utilization over time. It also emphasizes the importance of the ongoing assessment of telemedicine use with changing needs and preferences for eye care.

Telemedicine holds great potential in making eye care services accessible. However, the mere availability of telemedicine within a care setting is insufficient. It is equally important to address the challenges that hinder the effective utilization of telemedicine, particularly for marginalized populations, such as minority Americans, people living below the poverty line, rural residents, and the elderly. Failure to address these challenges not only limits telemedicine’s benefits, but also risks exacerbating existing disparities in eye care access and outcomes. In the U.S., many racial/ethnic minority groups are more likely to experience lower levels of digital and health literacy due to reasons such as acculturation and language proficiency.^9,42^ Data from the COVID-19 Research Database Consortium, which contains electronic medical records from all regions of the U.S., revealed that Hispanics were 41% less likely to engage in telehealth visits than non-Hispanic whites.^43^ Furthermore, even among minority populations who use telemedicine, many may be constrained to participate in telephone visits only rather than video visits due to their lack of suitable broadband and technology.^9,44-46^ Moreover, studies^47,48^ have found that households with lower incomes had lower telehealth usage, often attributable to barriers such as lack of broadband access.^9,42,46^ Access issues also exist for the elderly, a population that were found to be unready for video visits due to inexperience with using technology. Furthermore, older patients experience difficulties with hearing, communication, and dementia.^49^ Possible access accommodations include adding closed captioning or incorporating home visits for geriatric patients.^49^ Nonetheless, these disparities underscore the urgent need for future studies to address the potential disparities in access to telemedicine in eye care.

COVID-19 served as the tipping point for the adoption and awareness of telemedicine, as federal and state agencies as well as insurers adopted regulatory changes such as the lifting of telehealth restrictions and the issuing of telehealth waivers.^3^ The Kaiser Family Foundation found that temporary waivers issued in 49 states allowed out-of-state physicians to provide care for patients in a different state than where they were licensed to practice.^50^ Although these changes brought forth coverage and reimbursement benefits for patients, challenges still persisted in this area for physicians. Fortunately, the Telehealth Extension and Evaluation Act of 2022 allows for Medicare covered telehealth services to be active for up to two years after the public health emergency ends.^51^ This flexibility includes prescribing medications remotely, covering certain tests and equipment, and allowing rural health clinics and federally qualified health centers to serve patients regardless of location.^51^ To ensure the successful implementation of telemedicine in eye care in the long term, coverage needs to be extended in the future to allow for the continued reimbursement of eye care providers.

Many selected studies were conducted outside of the U.S., where the main form of health coverage is universal health care, typically through single-payer systems. A study conducted among oculoplastic surgeons from different countries reported that international physicians used telemedicine more often than physicians in the United States, at 72% and 47%, respectively.^17^ Despite these differences, the barriers to telemedicine adoption remained consistent, including low reimbursement rates, restrictive policies, and limited training programs.^52,53^ Furthermore, in the U.S., ophthalmology and optometry services are covered under separate insurance plans, and coverage may differ by insurance type (i.e., H.M.O., P.P.O., etc.) and across state lines, further complicating access to eye care via telemedicine. Although the studies in our review mainly focused on ophthalmology services, it is noteworthy that optometry services can also leverage telemedicine. A literature review by Massie et al.^54^ highlights the possibilities such as “comanagement” involving both an optometrist and ophthalmologist and standalone tele-optometry services.

## CONCLUSION

Our review demonstrates that telemedicine was well received by both patients and physicians for eye care during the pandemic, suggesting that telemedicine is a catalyst for potentially enhancing and restructuring eye care delivery. Addressing technological limitations, particularly in testing and imaging capabilities, is crucial for further improving the utilization and effectiveness of telemedicine in eye care. More research is required to identify populations and eye conditions/eye services that would benefit the most from telemedicine interventions.

Continued investigation is warranted to track evolving patient and physician perspectives toward telemedicine in the post-pandemic era.^55^ Despite the overall positive sentiment during the pandemic, significant organizational and systemic hurdles persist for the widespread adoption of telemedicine for eye care.^56^ COVID-19 transformed telemedicine utilization and the necessary changes put in place during the pandemic are imperative to ensure the sustained implementation of telemedicine in eye care.

## Data Availability

All data produced in the present study are available upon reasonable request to the authors.

